# Integrated treatment-decision algorithms for childhood TB: modelling diagnostic performance and costs

**DOI:** 10.1101/2025.06.20.25329945

**Authors:** Mary Gaeddert, Devan Jaganath, Abdulkadir Civan, Hoa Nguyen, Maryline Bonnet, Eric Wobudeya, Olivier Marcy, Manuela De Allegri, Claudia M. Denkinger

## Abstract

**Background:** To improve childhood tuberculosis (TB) diagnosis, treatment-decision algorithms (TDAs) with and without chest X-ray (CXR) were developed for children under age 10. We aimed to model diagnostic performance and costs of implementing TDAs in primary healthcare (PHC) and district hospital (DH) settings in Uganda.

**Methods:** We developed decision-tree models following the TDA pathway from evaluation to treatment-decision. We compared six scenarios with combinations of diagnostic testing (stool and respiratory Xpert, urine lipoarabinomannan, and/or CXR) at PHCs and DHs. Outcomes were diagnostic accuracy and cost per correct treatment-decision for a cohort of 10,000 children with presumptive TB using a Monte Carlo simulation from a health system perspective. Costs were reported in 2024 International dollars.

**Results:** In all scenarios, TDA’s had high sensitivity (80.8–91.9%) but low specificity (51.2-60.9%). Total diagnostic and treatment costs for the cohort were I$1,768,958–2,458,790; largely driven by overtreatment of false-positive cases. Diagnostic costs were mostly offset by reducing overtreatment. The cost per treatment-decision was lowest using mobile CXR at PHC (I$287.40) and highest with DH referral (I$445.84).

**Conclusion:** The TDAs have high sensitivity and can be implemented at PHCs with lower costs than DHs. Improving specificity and reducing treatment costs would enable affordable, large-scale implementation.

## INTRODUCTION

The burden of childhood tuberculosis (TB) remains high globally, driven by challenges in diagnosis and subsequent treatment initiation. There were an estimated 1.3 million new cases of TB and 191,000 deaths due to TB in children under 15 years of age in 2023,^1^ and it is estimated that 96% of deaths are in children not diagnosed and treated.^2^

In young children, TB often presents with non-specific symptoms and sputum-based testing is rarely feasible due to challenges obtaining samples.^3^ Even when sputum can be collected, sensitivity of culture and molecular testing is reduced due to the paucibacillary nature of childhood disease.^4^ Furthermore, chest X-ray (CXR) findings can be heterogeneous and difficult to interpret, especially in children living with HIV (CLHIV) and CXR is typically not available at primary health centers (PHC).^5^ Diagnostic capacity combining clinical assessment with clinical, radiographic, and laboratory information may only be present in district hospitals (DH). Decentralizing childhood TB services has been shown to increase case-finding and improve uptake at PHCs.^6^

To standardize diagnostic approaches and enable more children to initiate treatment earlier, two treatment-decision algorithms (TDAs) were developed for settings with and without CXR, guided by a large individual-patient data meta-analysis.^7^ The algorithms were designed with high sensitivity to reduce missed cases, at the expense of low specificity, potentially leading to significant overtreatment and associated costs. Previous studies evaluating the TDAs have only validated the clinical scoring system and have not incorporated other aspects such as different combinations of diagnostic tests and settings.^8, 9^ The World Health Organization (WHO) gave a conditional recommendation for the TDAs pending further validation of their accuracy and considerations for implementation, including costs, to guide programmatic adoption and scale-up.^10, 11^ The objectives of this analysis were to model the diagnostic performance and costs of implementing TDAs across different implementation scenarios in primary care and hospital-based settings.

## METHODS

### Study population and setting

We modelled the WHO TDAs among children under 10 years of age presenting with symptoms suggestive of pulmonary TB, including cough or fever for two weeks or more, poor appetite, weight loss or failure to thrive, and fatigue or reduced playfulness, who do not require urgent care. The setting for the analysis were outpatient clinics in PHCs and DHs in Uganda, which was chosen as a representative high-burden country with a TB incidence rate of 198/100,000 and 37% of TB cases are living with HIV.^1^ Of the TB cases reported nationally in 2023, over 12,000 (14%) were among children 0–14 years,^1^ but this may be underestimating the true burden.^12^ The current standard of care is screening children at health facility entry points, and evaluating those meeting criteria for presumptive TB with HIV testing, clinical examination, CXR, and collecting samples for bacteriological confirmation.^12^ In this setting, most resources for diagnostic testing, including X-ray facilities, on-site laboratories with GeneXpert, and clinical resources to perform gastric aspirate and sputum induction are at centralized facilities. However, approximately 54% of children initially present at PHCs, which have limited access to diagnostics, so children with presumptive TB are often referred to DHs.^13^

### WHO Treatment Decision Algorithms

Each TDA follows a sequence of evaluations. Because the algorithms aim to detect as many cases as possible, a negative result on any step leads to further assessment until a treatment decision is reached. Children at high-risk for rapid disease progression (under 2 years of age, living with HIV, or with severe acute malnutrition) are tested for TB immediately, including molecular testing on stool or respiratory samples with GeneXpert, and urine lateral flow lipoarabinomannan (LF-LAM, Determine TB LAM, Abbott, Chicago) for CLHIV. The children not in a high-risk group are assessed for other likely conditions related to their symptoms and return for follow-up in two weeks. Children whose symptoms resolve by the follow-up visit exit the algorithm and undergo no further testing. Children with persistent symptoms continue for molecular testing. If these tests are negative or not available, the child proceeds to the clinical scoring step, which considers history of TB contact and presence of individual symptoms. Algorithm A (TDA-A) includes CXR findings in the scoring and can be used where X-ray is available. While this is typically relevant for DHs, mobile vans with portable X-ray machines may be available to expand access to PHCs.^14^ Algorithm B (TDA-B) is for settings without X-ray, typically PHCs, and only includes clinical signs and symptoms.

### Model structure

We developed six scenarios comparing a range of strategies at PHC and the centralized DH approach. We converted the TDAs into a decision-tree model with separate arms for each scenario, following the above-described pathway from initial evaluation to treatment decision (Figure 1). Scenario 1 (‘TDA-B’) considered the simplest scenario with only clinical diagnosis using the TDA-B scoring system and no molecular testing. Scenario 2 (‘TDA-A’) explored the impact of adding mobile CXR and use of the TDA-A scoring. Scenario 3 (‘Stool + TDA-B’) and Scenario 4 (‘Stool + TDA-A’) included molecular testing with Xpert Ultra on stool samples and urine LF-LAM for CLHIV, with or without mobile CXR. Scenario 5 (‘Stool + TDA-B + Referral’) began with stool testing at PHC and referred a portion of children with a negative result to DHs for respiratory Xpert Ultra testing and TDA-A with CXR. Children in the high-risk group were more likely to be referred, and those who remained at PHC were evaluated with the TDA-B score. This scenario most closely reflected the current standard of care in Uganda. Scenario 6 (‘Referral’) reflected the centralized approach where all children had CXR and respiratory sample testing at DHs.

**Figure 1.**
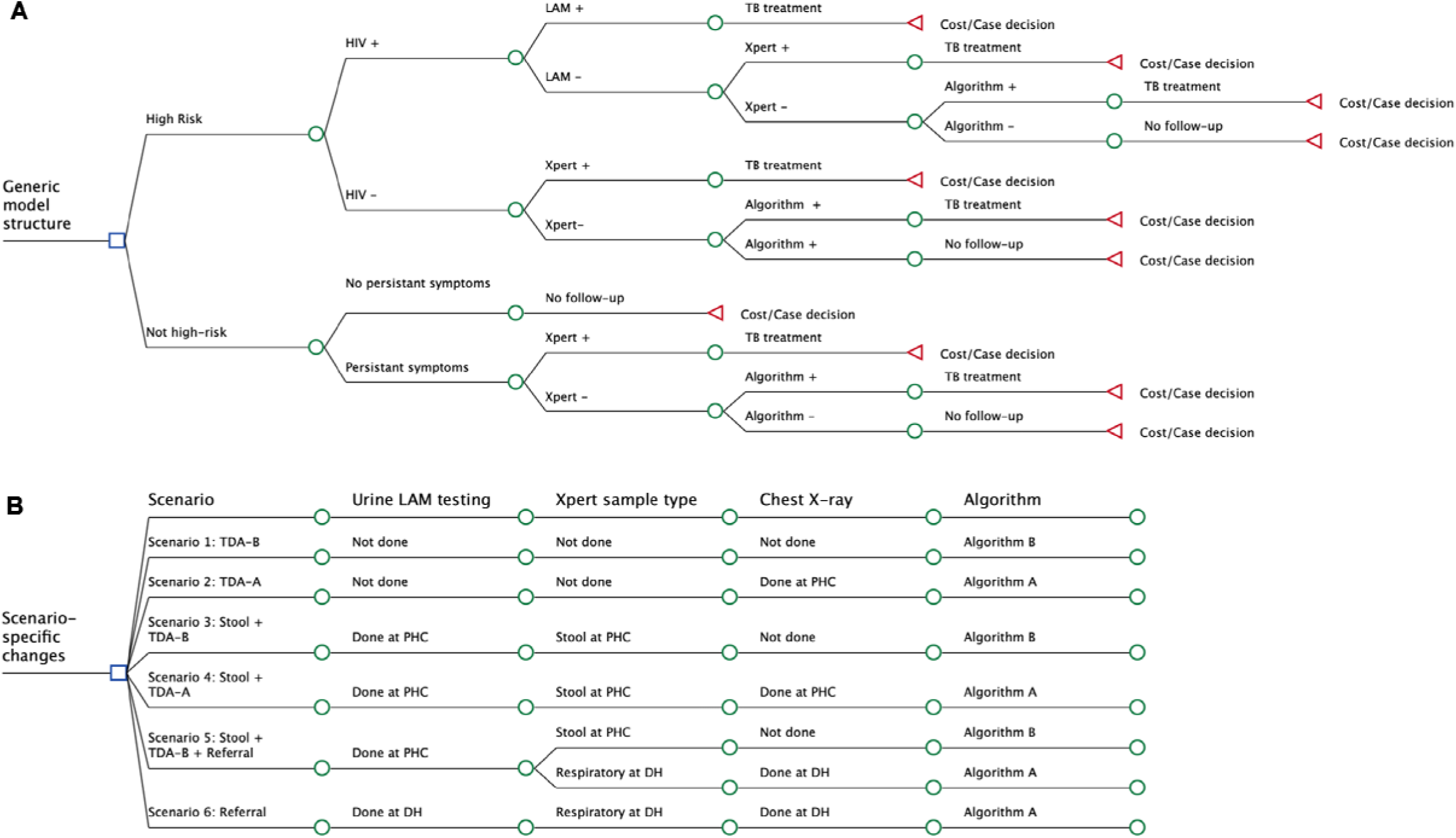
Decision-tree model and scenario-specific changes. Part A shows a simplified version of the clinical pathway converted into a decision-tree model for each scenario. Part B shows the diagnostic tests used for each model scenario.

### Model parameters and analysis

Estimates for clinical and cost parameters matching the modelled setting were obtained from the literature (Table S1). Several clinical parameter estimates were obtained from a multi-site study on decentralization of TB testing and the authors (MB, OM, EW) provided additional data for the Ugandan site.^13^ We also consulted expert opinion (see Acknowledgement) for input on clinical parameters, including TB prevalence and resolution of symptoms during follow-up. Due to limited data available on pre-diagnostic loss and the possible impact of new strategies for children in this setting, loss to follow-up before and during the diagnostic process was not included. The costs of clinical examination, sample collection, HIV testing, and TB molecular testing were included as specified for each scenario. The cost of respiratory sample collection at DH included gastric aspirates for children under five and induced or expectorated sputum for older children. TB treatment costs included direct costs of medications and follow-up visits until completion of therapy. Patient costs such as transportation and the burden of treatment (i.e., giving daily medications and side effects) were not included. Drug-resistant TB was not included as the rates are low among children in Uganda.^15^

The outcomes were diagnostic accuracy and cost per correct treatment-decision (both true-positives and true-negatives). Treatment costs were reported separately for true TB cases, overtreatment, and risk group. Outcomes were calculated for a cohort of 10,000 children using a Monte Carlo simulation. The analysis adopted a health system perspective, using a time horizon of one year for program implementation and no discounting was applied. Costs were converted to 2024 International dollars (I$) using World Bank inflation data.^16, 17^

One-way and probabilistic sensitivity analyses (PSA) were conducted to evaluate how uncertainty in model parameters impacted outcomes. TreeAge Pro 2024 was used for analysis. Details of the evaluation are reported following the Consolidated Health Economic Evaluation Reporting Standards (CHEERS) guidance (Table S2).^18^ Ethical approval was not required as all parameters were obtained from published literature and there was no human subject participation.

## RESULTS

### Diagnostic performance

Overall, the diagnostic accuracy of the scenarios was moderate, ranging from 55.0–61.6%, balancing a high sensitivity and low specificity (Table 1, full results in Table S3). The sensitivity was high in all scenarios, ranging from 80.8–91.9%, indicating that the TDAs are not missing many TB cases. However, the specificity was consistently low (51.2–60.9%), resulting in a low positive predictive value (PPV) (5.5–6.3%), particularly in Scenarios 1-5 at PHCs with low prevalence of TB disease (3%). The negative predictive value (NPV) was above 98% across all scenarios.

**Table 1.** Diagnostic accuracy of scenarios for a cohort of 10,000 children under 10 years of age with presumptive TB.

| Scenario* | Scenario 1:<br>TDA-B | Scenario 2:<br>TDA-A | Scenario 3:<br>Stool + TDA-B | Scenario 4:<br>Stool + TDA-A | Scenario 5:<br>Stool + TDA-B +<br>Referral | Scenario 6:<br>Referral |
| --- | --- | --- | --- | --- | --- | --- |
| TDA | Algorithm B | Algorithm A | Algorithm B | Algorithm A | Algorithm A and B | Algorithm A |
| Chest X-ray Available | No | Yes | No | Yes | Partial | Yes |
| TB-specific testing | No** | No** | Yes | Yes | Yes | Yes |
| Accuracy, overall | 56.8% | 61.6% | 55.0% | 59.6% | 57.6% | 55.2% |
| High-risk group | 32.9% | 40.3% | 30.0% | 37.3% | 34.2% | 41.6% |
| Not high-risk group | 86.0% | 87.4% | 85.4% | 86.7% | 86.1% | 86.4% |
| Sensitivity, overall | 80.8% | 82.7% | 86.3% | 87.0% | 86.3% | 91.9% |
| High-risk group | 86.7% | 89.8% | 92.8% | 94.0% | 93.4% | 97.0% |
| Not high-risk group | 73.8% | 74.5% | 78.7% | 78.7% | 78.0% | 79.2% |
| Specificity, overall | 56.1% | 60.9% | 54.0% | 58.7% | 56.6% | 51.2% |
| High-risk group | 31.2% | 38.8% | 28.0% | 35.5% | 32.3% | 35.6% |
| Not high-risk group | 86.4% | 87.8% | 85.6% | 86.9% | 86.3% | 87.1% |
| Positive-predictive value | 5.5% | 6.3% | 5.6% | 6.3% | 5.9% | 16.8% |
| High-risk group | 3.8% | 4.4% | 3.9% | 4.3% | 4.1% | 14.2% |
| Not high-risk group | 14.9% | 16.5% | 15.0% | 16.3% | 15.6% | 38.0% |
| Negative predictive value | 98.9% | 99.1% | 99.2% | 99.3% | 99.2% | 98.3% |
| High-risk group | 98.7% | 99.2% | 99.2% | 99.5% | 99.4% | 99.1% |
| Not high-risk group | 99.0% | 99.1% | 99.2% | 99.2% | 99.2% | 97.7% |
| Evaluated with clinical score | 64.8% | 64.8% | 62.1% | 62.1% | 61.1% | 67.3% |
| High-risk group | 100% | 100% | 96.2% | 96.2% | 94.8% | 88.1% |
| Not high-risk group | 21.9% | 21.9% | 20.4% | 20.4% | 20.1% | 19.3% |

\*The setting for all scenarios is at primary health clinics, unless specified as referral to district hospitals
\*\*Scenario 1 and Scenario 2 follows the algorithm without TB-specific testing (Xpert Ultra or LF-LAM)
Legend for abbreviations: urine lateral flow lipoarabinomannan (LF-LAM), Tuberculosis (TB), treatment-decision algorithm (TDA)

**Table 2.**
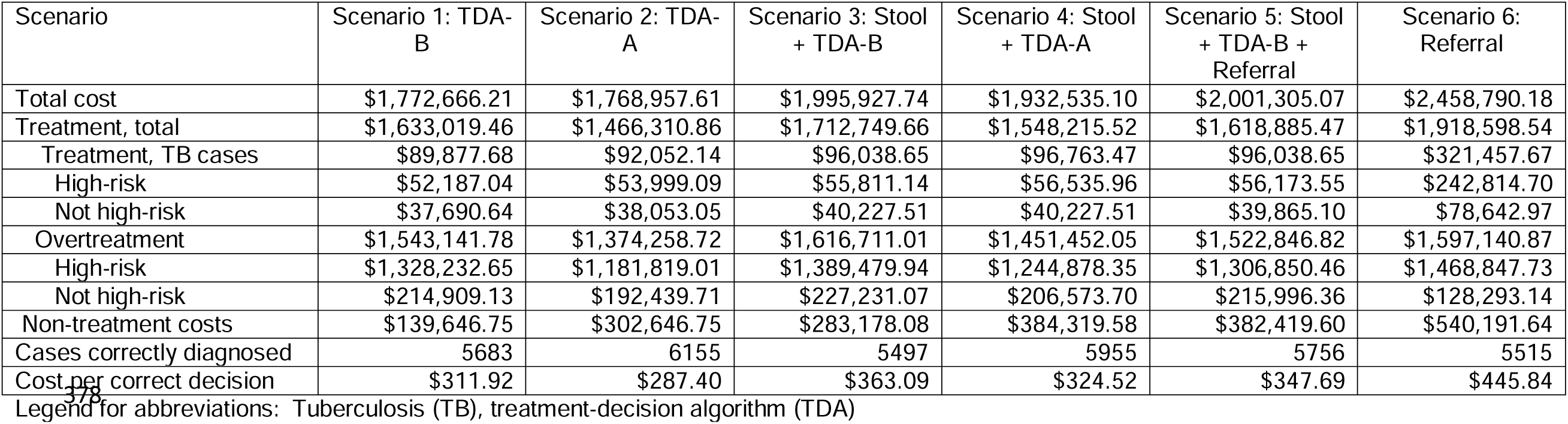
Costs of diagnostic testing and TB treatment of scenarios for a cohort of 10,000 children under 10 years of age with presumptive TB.

When comparing Scenario 1 and 2 (clinical diagnosis with and without CXR respectively), the addition of mobile CXR at PHCs improved both sensitivity (80.8% versus 82.7%, respectively) and specificity (56.1% versus 60.9%, respectively). The addition of molecular testing, with or without CXR, in Models 3 and 4 improved sensitivity (86.3% versus 87.0%, respectively) and specificity (54.0 versus 58.7%, respectively) compared to Scenarios 1 and 2. Referring either some or all children for CXR and respiratory sample testing (Scenarios 5 and 6, respectively) improved sensitivity (86.3% versus 91.9%, respectively) but lowered specificity (56.6% versus 51.2%, respectively). These relationships were similar when comparing high and low-risk groups. However, low-risk groups had on average 10% lower sensitivity and 30% higher specificity compared to the overall results.

### Costs

The total costs of TB testing and treatment for a cohort of 10,000 children ranged from I$1,768,958 in Scenario 2 to I$2,458,790 in Scenario 6 (Table 3). The costs were driven by the overtreatment of false-positive cases, which resulted in more than I$1.4 million in every scenario. The lower specificity in the high-risk group resulted in a larger proportion of overtreatment costs than in the low-risk group. Due to the low prevalence of TB at PHC, the cost for treating true TB cases was only 5% of total costs for Scenarios 1-5.

The diagnostic costs are lowest in Scenario 1 with only clinical diagnosis (I$139,647) and highest in Scenario 6 with referral testing (I$540,192). When comparing scenarios, the increased cost of diagnostics was mostly offset by the decreased cost of overtreatment. For example, the addition of mobile CXR between Scenarios 3 and 4 increased diagnostic costs by I$101,142 but reduced overtreatment by I$165,259. However, referring all children in Scenario 6 substantially increased diagnostic costs but did not reduce overtreatment. This is reflected in the cost per correct treatment-decision, which was lowest using mobile CXR at PHC (Scenario 2, I$287.40) and highest with DH referral (Scenario 6, I$445.84).

### Sensitivity Analyses

One-way sensitivity analyses showed that the parameters with the greatest impact were the specificity of TDA’s, cost of TB treatment, and proportion of children who were high-risk (Figure 2). As TDA specificity increases, reduction in overtreatment lowers the cost per case. Decreasing treatment costs and proportions of high-risk children also decreased the cost per case. The PSA indicated Scenario 2 was more cost-effective than other scenarios across a range of willingness-to-pay thresholds (Figure 3). Additional PSAs are in Figure S1.

**Figure 2.**
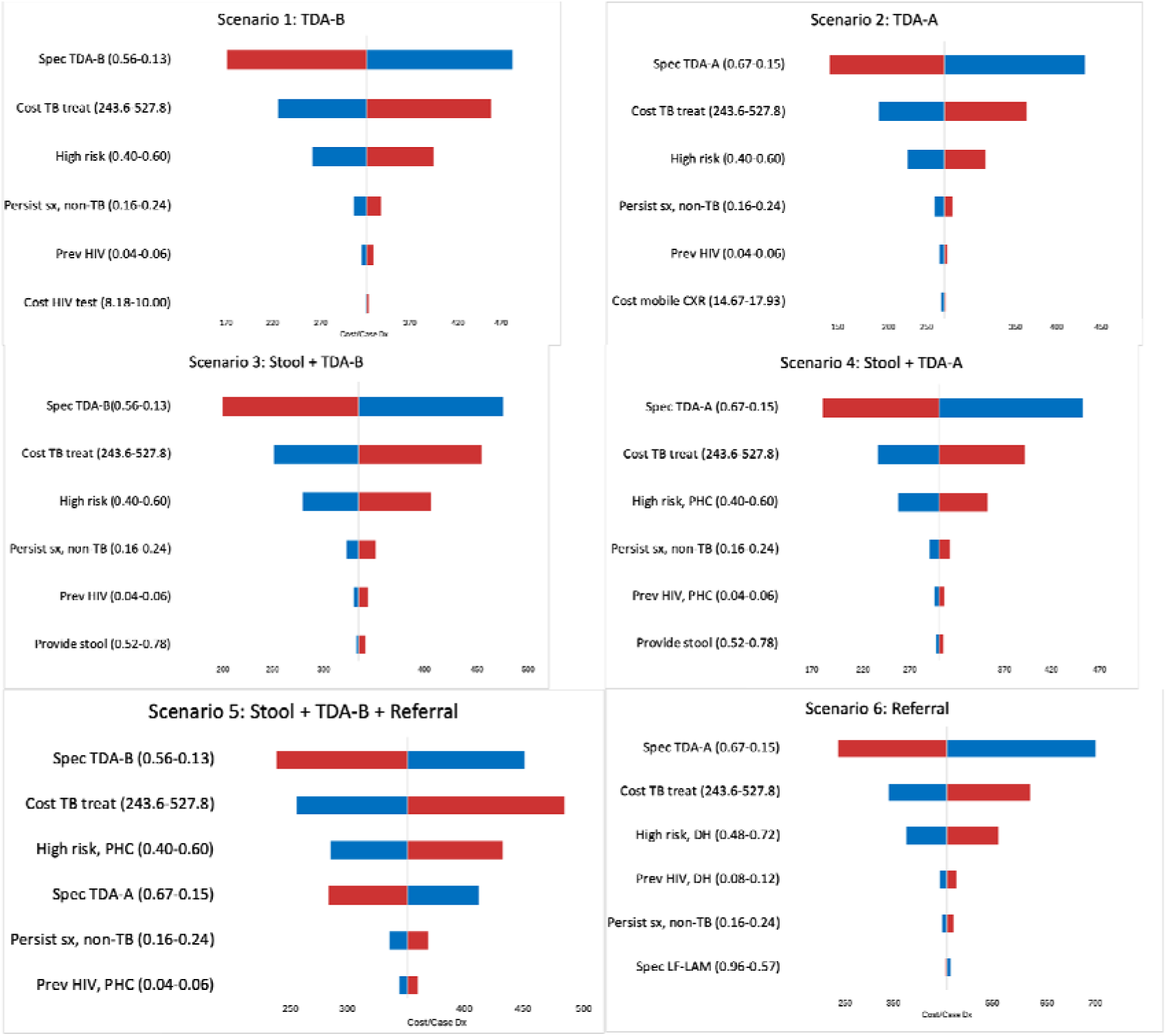
One-way sensitivity analyses for each scenario. Parameters are shown in order of decreasing impact on the outcome of cost per correct treatment-decision. Blue=low range of parameter, red=high range of parameter

**Figure 3:**
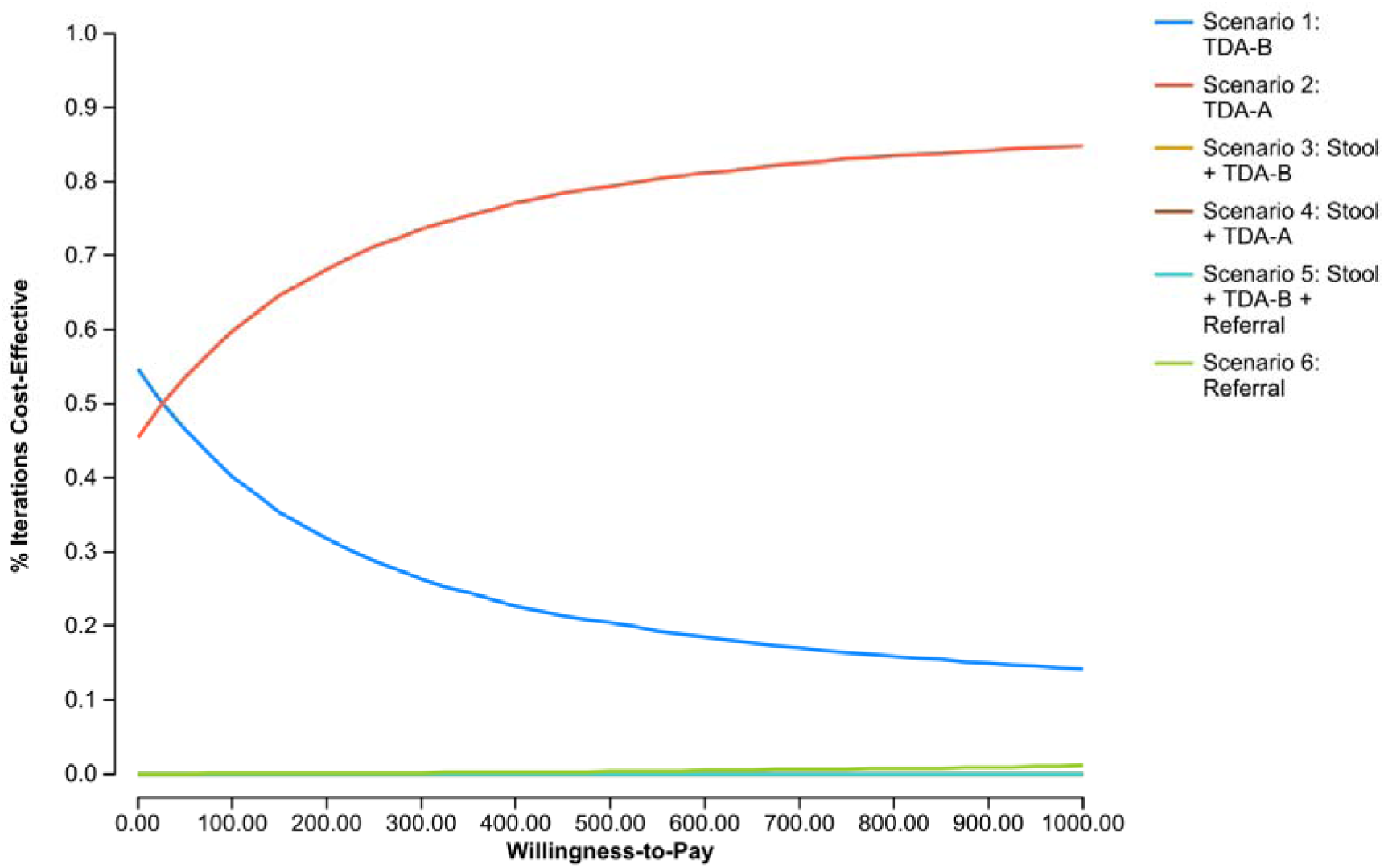
Cost-Effectiveness Acceptability Curve. Probabilistic sensitivity analysis for the joint uncertainty of all parameters using a Monte-Carlo simulation with 10,000 iterations. The cost-effectiveness acceptability curve shows which scenario was most cost-effective at the given willingness-to-pay threshold.

## DISCUSSION

This is the first analysis to model both the diagnostic accuracy and costs of the new TDAs in a cohort of children across a range of implementation scenarios. The TDAs were developed to support the clinical assessment for childhood TB and reduce the diagnostic gap. We showed that the TDAs have high sensitivity to detect TB cases with high NPV, but their low specificity leads to substantial overtreatment costs. The current standard of referring children to DHs had both the highest sensitivity and highest cost. However, expanding diagnostic capacity at PHCs improved specificity, reduced overtreatment, and consequently offset costs. These findings suggest that implementation of TDAs at PHCs with and without TB-specific testing would have high sensitivity to detect TB, and lower costs than referral to a DH, but further improvements are needed to reduce costs of overtreatment.

The scenarios with DH referral do not substantially improve accuracy or reduce costs compared to scenarios at PHCs. Although children have access to respiratory testing and CXR at DHs, if these results are negative then children could still be initiated on TB treatment based on the less-specific clinical score. At the same time, there are additional costs for the DH assessment and any previous PHC visits before referral. In PHC scenarios, mobile CXR improved specificity and higher diagnostic costs were offset by the reduction in overtreatment. Stool-based testing did not improve accuracy, again as children with negative stool tests would then be assessed with the clinical score. The difference in clinical pathway for high-risk children increased the sensitivity, but with a trade-off of lower specificity and associated higher overtreatment cost. However, it is important to recognize the benefits of microbiological confirmation, including detection of drug resistance.^19^ CXR also has benefits of classifying disease severity and eligibility for shorter treatment regimens, or identifying alternative diagnoses.^11^ Additional benefits of decentralization for children and caregivers include reduced costs and burden of visits to referral facilities.

However, it is important to recognize the high total costs. Our model estimated the costs for a cohort of 10,000 children, including diagnostics and treatment, were over I$1.7 million for all scenarios. The 2023 funding for TB in Uganda was $32 million and 84% came from international sources.^1^ In a time of decreased global health funding, it may not be feasible for national programs or donors to cover costs of expanding services, and the cost of diagnostics alone may not be affordable for many countries.^20^ An analysis of decentralization strategies, which included scale-up costs such as training and equipment, found that decentralization to the PHC level would likely not be cost-effective,^21^ and a cost-effectiveness analysis of TB screening in Uganda found similar challenges in an adult population with low TB prevalence.^22^ Our sensitivity analyses showed that improved TDA specificity, especially among high-risk children, and reduced treatment costs would provide the greatest impact. Moreover, implementing newly-recommended shorter treatment regimens for children with non-severe disease would also reduce treatment costs. ^10^ ^23^

Strengths of this analysis include using parameters from studies conducted in similar high-burden settings, including a childhood TB the decentralization study in Uganda.^12^ We also consulted expert opinion to support the limited data available for some clinical parameters. However, there are still limited data available for childhood TB, and the impact of this uncertainty was explored in the sensitivity analyses. These models did not include pre-diagnostic loss to follow-up, so our estimations are likely overoptimistic. We also did not include patient costs which would likely support more patient-centered algorithms at PHCs. Implementation will require additional resources for training and supporting healthcare workers.

Clinical studies to validate TDA performance in high-burden settings are ongoing and the results will inform future implementation. When these studies are completed, it will be valuable to conduct formal budget impact analyses. Additional evaluations including patient costs, caregiver preferences regarding location of care, and feedback from healthcare workers on their experience using the algorithms will inform stakeholder decision-making. Modifications to improve algorithm performance, especially increasing specificity (e.g. through more scalable and accessible pathogen-based diagnostics) should be considered.

## CONCLUSIONS

Increasing children’s access to TB diagnostic tools is important. Our models indicate that the TDAs have high sensitivity and negative predictive value, enabling increased detection of childhood TB, and can be implemented at primary care centers at lower cost than district hospitals. However, the low specificity and subsequent overtreatment costs could reduce the feasibility of implementation in real-world settings, unless there are further efforts to improve specificity and reduce treatment costs.

## Data Availability

All data produced in the present work are contained in the manuscript.

## List of abbreviations

CLHIV: Children living with HIV
CXR: Chest X-ray
DH: District hospital
LF-LAM: Lateral flow urine lipoarabinomannan assay
NPV: negative predictive value
PHC: Primary health center
PPV: positive predictive value
PSA: probabilistic sensitivity analysis
TB: Tuberculosis
TDA: treatment-decision algorithm
WHO: World Health Organization

## Acknowledgements

We would like to thank the following experts for providing input on the clinical parameters for childhood TB: Beate Kampmann, Ben Marais, and Steve Graham. We would also like to thank Ken Gunasekera and James Seddon for providing input on how the algorithms were originally developed.

This work was supported by the National Institute of Allergy and Infectious Diseases at the National Institutes of Health [U01AI152087 to CMD]; the National Heart, Lung, and Blood Institute at the National Institutes of Health [K23HL153581 to DJ]; and the German Center for Infection Research (DZIF) [TTU.02.813, funding indicator 8029802813 to MG]. The funders had no role in the identification, design, conduct, and reporting of the analysis. The authors have no conflicts of interest to report.

This analysis used data from published studies and did not require ethical approval.

## Notes

### Competing Interest Statement

The authors have declared no competing interest.

