## Supplemental materials for "Integrated treatment-decision algorithms for childhood TB: modelling diagnostic performance and costs"

**Table S1:** Model parameter estimates for children under 10 years of age with presumptive TB in Uganda

**Table S2:** Consolidated Health Economic Evaluation Reporting Standards checklist

**Table S3:** 2x2 tables with diagnostic accuracy results for cohort of 10,000 children with presumptive TB under 10 years of age

**Figure S1:** Additional Probabilistic Sensitivity Analyses

| **Table S1.** Model parameter estimates for children under 10 years of age with presumptive TB in Uganda | | | | |
| --- | --- | --- | --- | --- |
| Clinical Parameters | Base Case | Range | Distribution | Reference |
| Prevalence of TB, PHC | 0.03 | 0.02–0.04 | Beta | (1) |
| Prevalence of TB, DH | 0.10 | 0.08–0.12 | Beta | (1) |
| Prevalence of HIV, PHC | 0.05 | 0.04–0.06 | Beta | (2) |
| Prevalence of HIV, DH | 0.10 | 0.08–0.12 | Beta | (2) |
| Proportion high-risk, PHC | 0.50 | 0.40–0.60 | Beta | (1) |
| Proportion high-risk, DH | 0.60 | 0.48–0.72 | Beta | (1) |
| Referral to DH, high risk | 0.35 | 0.28-0.42 | Beta | (1) |
| Referral to DH, low risk | 0.20 | 0.16–0.24 | Beta | (1) |
| Persistent symptoms, TB positive | 0.80 | 0.64–0.96 | Beta | (3, 4) |
| Persistent symptoms, TB negative | 0.20 | 0.16–0.24 | Beta | (3, 4) |
| Sensitivity of respiratory Xpert Ultra, children without HIV | 0.73 | 0.65–0.80 | Beta | (5-7) |
| Specificity of respiratory Xpert Ultra, children without HIV | 0.97 | 0.96–0.98 | Beta | (5-7) |
| Sensitivity of respiratory Xpert Ultra, children with HIV | 0.64 | 0.44–0.80 | Beta | (5-7) |
| Specificity of respiratory Xpert Ultra, children with HIV | 0.98 | 0.93–1.0 | Beta | (5-7) |
| Sensitivity of stool Xpert Ultra | 0.77 | 0.66–0.85 | Beta | (8) |
| Specificity of stool Xpert Ultra | 0.98 | 0.96–0.99 | Beta | (8) |
| Sensitivity of urine LAM, children with HIV | 0.47 | 0.33–0.60 | Beta | (9) |
| Specificity of urine LAM, children with HIV | 0.76 | 0.57–0.96 | Beta | (9) |
| Sensitivity of TDA-A with CXR | 0.88 | 0.71–0.95 | Beta | (10) |
| Specificity of TDA-A with CXR | 0.37 | 0.15–0.67 | Beta | (10) |
| Sensitivity of TDA-B | 0.86 | 0.68–0.94 | Beta | (10) |
| Specificity of TDA-B | 0.30 | 0.13–0.56 | Beta | (10) |
| Stool sample available | 0.65 | 0.52–0.78 | Beta | (1) |
| Urine sample available | 0.65 | 0.52–0.78 | Beta | (11) |
| Cost Parameters, I$ | Base Case | Range | Distribution | Reference |
| Outpatient visit, PHC | $3.25 | 2.92–3.57 | Gamma | (12) |
| Outpatient visit, DH | $4.56 | 4.11–5.02 | Gamma | (12) |
| HIV testing | $9.09 | 8.18–10.00 | Gamma | (13) |
| Urine LAM testing | $4.90 | 4.41–5.39 | Gamma | (14) |
| Chest X-ray, DH | $11.37 | 10.23–12.50 | Gamma | (15) |
| Mobile Chest X-ray, PHC | $16.30 | 14.67–17.93 | Gamma | (16) |
| Stool collection | $1.99 | 1.79–2.19 | Gamma | (17) |
| Expectorated sputum | $1.88 | 1.69–2.07 | Gamma | (17) |
| Induced sputum | $27.57 | 24.81–30.32 | Gamma | (17) |
| Gastric aspirate | $5.21 | 4.69–5.73 | Gamma | (17) |
| Sample transport | $1.63 | 1.47–1.79 | Gamma | (18) |
| Stool processing | $4.54 | 4.08–4.99 | Gamma | (19) |
| Xpert Ultra testing | $21.86 | 19.67–24.04 | Gamma | (18) |
| Cost of TB treatment, 6 months | $362.41 | 243.61–527.82 | Gamma | (20) |
| Legend for abbreviations: chest X-ray (CXR), district hospital (DH), Human immunodeficiency virus (HIV), International dollars (I$), urine lateral flow lipoarabinomannan (LF-LAM), primary health center (PHC), Tuberculosis (TB), treatment-decision algorithm (TDA) | | | | |

**Table S2:** Consolidated Health Economic Evaluation Reporting Standards checklist

| **Topic** | **No.** | **Item** | **Location where item is reported** |
| --- | --- | --- | --- |
| **Title** |  |  |  |
|  | 1 | Identify the study as an economic evaluation and specify the interventions being compared. | Title page |
| **Abstract** |  |  |  |
|  | 2 | Provide a structured summary that highlights context, key methods, results, and alternative analyses. | Abstract |
| **Introduction** |  |  |  |
| **Background and objectives** | 3 | Give the context for the study, the study question, and its practical relevance for decision making in policy or practice. | Paragraph 2-3 |
| **Methods** |  |  |  |
| **Health economic analysis plan** | 4 | Indicate whether a health economic analysis plan was developed and where available. | Not done |
| **Study population** | 5 | Describe characteristics of the study population (such as age range, demographics, socioeconomic, or clinical characteristics). | Methods, Paragraph 1 |
| **Setting and location** | 6 | Provide relevant contextual information that may influence findings. | Methods, Paragraph 1 |
| **Comparators** | 7 | Describe the interventions or strategies being compared and why chosen. | Methods, Paragraphs 2-3 |
| **Perspective** | 8 | State the perspective(s) adopted by the study and why chosen. | Methods, Pararaph 5 |
| **Time horizon** | 9 | State the time horizon for the study and why appropriate. | Methods, Paragraph 5 |
| **Discount rate** | 10 | Report the discount rate(s) and reason chosen. | Methods, Paragraph 5 |
| **Selection of outcomes** | 11 | Describe what outcomes were used as the measure(s) of benefit(s) and harm(s). | Methods, Paragraph 5 |
| **Measurement of outcomes** | 12 | Describe how outcomes used to capture benefit(s) and harm(s) were measured. | Methods, Paragraph 5 |
| **Valuation of outcomes** | 13 | Describe the population and methods used to measure and value outcomes. | Methods, Paragraph 5 |
| **Measurement and valuation of resources and costs** | 14 | Describe how costs were valued. | Methods, Paragraph 5 |
| **Currency, price date, and conversion** | 15 | Report the dates of the estimated resource quantities and unit costs, plus the currency and year of conversion. | Methods, Paragraph 5 |
| **Rationale and description of model** | 16 | If modelling is used, describe in detail and why used. Report if the model is publicly available and where it can be accessed. | Methods, Paragraph 3, Figure 1 |
| **Analytics and assumptions** | 17 | Describe any methods for analysing or statistically transforming data, any extrapolation methods, and approaches for validating any model used. | Methods, Paragraph 4-6 |
| **Characterising heterogeneity** | 18 | Describe any methods used for estimating how the results of the study vary for subgroups. | Methods, Paragraph 5 |
| **Characterising distributional effects** | 19 | Describe how impacts are distributed across different individuals or adjustments made to reflect priority populations. | Not done |
| **Characterising uncertainty** | 20 | Describe methods to characterise any sources of uncertainty in the analysis. | Methods, Paragraph 6 |
| **Approach to engagement with patients and others affected by the study** | 21 | Describe any approaches to engage patients or service recipients, the general public, communities, or stakeholders (such as clinicians or payers) in the design of the study. | Not done |
| **Results** |  |  |  |
| **Study parameters** | 22 | Report all analytic inputs (such as values, ranges, references) including uncertainty or distributional assumptions. | Supplemental Table 1 |
| **Summary of main results** | 23 | Report the mean values for the main categories of costs and outcomes of interest and summarise them in the most appropriate overall measure. | Results, Tables 2-3 |
| **Effect of uncertainty** | 24 | Describe how uncertainty about analytic judgments, inputs, or projections affect findings. Report the effect of choice of discount rate and time horizon, if applicable. | Results, Figure 2, Figure 3, Supplemental Figure 1 |
| **Effect of engagement with patients and others affected by the study** | 25 | Report on any difference patient/service recipient, general public, community, or stakeholder involvement made to the approach or findings of the study | Not applicable |
| **Discussion** |  |  |  |
| **Study findings, limitations, generalisability, and current knowledge** | 26 | Report key findings, limitations, ethical or equity considerations not captured, and how these could affect patients, policy, or practice. | Discussion |
| **Other relevant information** |  |  |  |
| **Source of funding** | 27 | Describe how the study was funded and any role of the funder in the identification, design, conduct, and reporting of the analysis | Acknowledgements, Funding |
| **Conflicts of interest** | 28 | Report authors conflicts of interest according to journal or International Committee of Medical Journal Editors requirements. | Acknowledgements, COI |

**Table S3. Diagnostic accuracy results for a cohort of 10,000 children under 10 years of age with presumptive TB**

|  | Overall | | | High-risk | | | Not high-risk | | |
| --- | --- | --- | --- | --- | --- | --- | --- | --- | --- |
| Scenario 1: TDA-B | | | | | | | | | |
|  | TB+ | TB- |  | TB+ | TB- |  | TB+ | TB- |  |
| Algorithm + | 248 | 4258 | 4506 | 144 | 3665 | 3809 | 104 | 593 | 697 |
| Algorithm - | 59 | 5435 | 5494 | 22 | 1663 | 1685 | 37 | 3772 | 3809 |
|  | 307 | 9693 | 10000 | 166 | 5328 | 5494 | 141 | 4365 | 4506 |
| Scenario 2: TDA-A | | | | | | | | | |
| Algorithm + | 254 | 3792 | 4046 | 149 | 3261 | 3410 | 105 | 531 | 636 |
| Algorithm - | 53 | 5901 | 5954 | 17 | 2067 | 2084 | 36 | 3834 | 3870 |
|  | 307 | 9693 | 10000 | 166 | 5328 | 5494 | 141 | 4365 | 4506 |
| Scenario 3: Stool + TDA-B | | | | | | | | | |
| Algorithm + | 265 | 4461 | 4726 | 154 | 3834 | 3988 | 111 | 627 | 738 |
| Algorithm - | 42 | 5232 | 5232 | 12 | 1494 | 1506 | 30 | 3738 | 3768 |
|  | 307 | 9693 | 10000 | 166 | 5328 | 5494 | 141 | 4365 | 4506 |
| Scenario 4: Stool + TDA-A | | | | | | | | | |
| Algorithm + | 267 | 4005 | 4272 | 156 | 3435 | 3591 | 111 | 570 | 681 |
| Algorithm - | 40 | 5688 | 5728 | 10 | 1893 | 1903 | 30 | 3795 | 3825 |
|  | 307 | 9693 | 10000 | 166 | 5328 | 5494 | 141 | 4365 | 4506 |
| Scenario 5: Stool + TDA-B + Referral | | | | | | | | | |
| Algorithm + | 265 | 4202 | 4467 | 155 | 3606 | 3761 | 110 | 596 | 706 |
| Algorithm - | 42 | 5491 | 5533 | 11 | 1722 | 1733 | 31 | 3769 | 3800 |
|  | 307 | 9693 | 10000 | 166 | 5328 | 5494 | 141 | 4365 | 4506 |
| Scenario 6: Referral | | | | | | | | | |
| Algorithm + | 887 | 4407 | 5294 | 670 | 4053 | 4723 | 217 | 354 | 571 |
| Algorithm - | 78 | 4628 | 4706 | 21 | 2236 | 2257 | 57 | 2392 | 2449 |
|  | 965 | 9035 | 10000 | 691 | 6289 | 6980 | 274 | 2746 | 3020 |
| Legend for abbreviations: Tuberculosis (TB), treatment-decision algorithm (TDA) | | | | | | | | | |

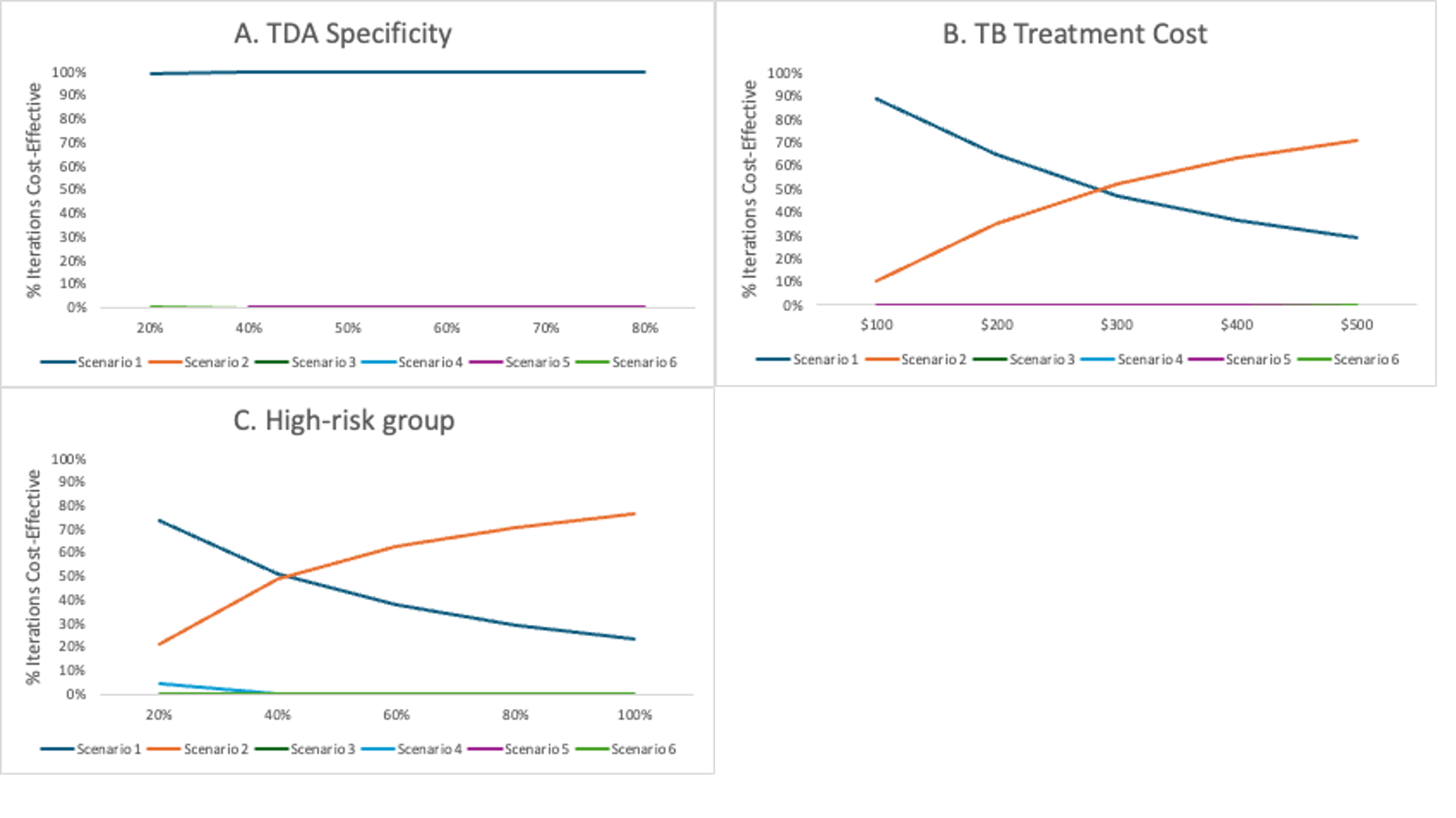

**Figure S1: Additional Probabilistic Sensitivity Analyses**

Probabilistic sensitivity analysis for the joint uncertainty of parameters using a Monte-Carlo simulation with 10,000 iterations. The parameter value of interest is fixed as specified in each figure and the range of all other parameters are varied. The cost-effectiveness acceptability curve shows which scenario was most cost-effective at the given willingness-to-pay threshold.

Scenario 1= dark blue, Scenario 2= orange, all remaining scenarios are at or close to 0%.
